# Illustration of a Novel Gut-Brain Axis of Alcohol Withdrawal, Withdrawal-Associated Depression, Craving and Alcohol-Severity Index in Alcohol Use Disorder Patients

**DOI:** 10.1101/2022.05.15.22275115

**Authors:** Vatsalya Vatsalya, Ranganathan Parthasarathy, Joris Verster, Amor C. Royer, Manasa Sagaram, Zarlakhta Zamani, Huirong Hu, Melanie L. Schwandt, Leggio Lorenzo, Maiying Kong, Vijay A Ramchandani, Wenke Feng, Xiang Zhang, Craig J. McClain

**Author notes:** **Corresponding Author** Vatsalya Vatsalya MD PgD MSc MS, Department of Medicine, University of Louisville School of Medicine, 505 S. Hancock St., CTR Room 514A, Louisville, KY 40202. **Author contribution:** VV is the project PI and designed the study. VV, MLS, HH, ZZ, and MK participated in the clinical sample and data analyses. VV, RP, AR, MS, MK, MS, WF, VAR, XZ, CJM, and JV interpreted the results. VV, RP, AR, JV, ZZ and CJM wrote the manuscript. CJM, MLS, MK, VAR, JV, WF, XZ and VV critically reviewed the manuscript and contributed scientifically. All authors have approved the submission version of this manuscript. **Trial registration:** ClinicalTrials.gov identifier – NCT# 00106106. **Ethics approval and consent to participate:** The study was approved by the site/s’ Institutional Review Board(s) (Ethics committee). All patients included in this study consented to participate before the beginning of the study. **Proprietorship:** This article is a work of the University of Louisville Alcohol Research Center and the National Institutes of Health. This manuscript is in the public domain in the USA. **Availability of data and materials:** The datasets analyzed during the current study are available from the corresponding author on reasonable request. Email address for the readers to contact the author to obtain the data. **Disclosure:** This article has not been published, nor is it under consideration for publication elsewhere at the time of submission. **Reprint requests:** Vatsalya Vatsalya MD, PgD, MSc, MS. Department of Medicine, University of Louisville School of Medicine; 505 S. Hancock St., CTR Room 514A, Louisville KY 40202 USA.

## Abstract

Pathways underlying the gut-brain axis and pro-inflammatory cytokine production influence brain functions and behavior. Alcohol use disorder (AUD) patients exhibit domains such as alcohol withdrawal, depression, and craving; and the gut-immune response may play a significant role in these domains of AUD. This study examined the role of intestinal permeability, pro-inflammatory cytokines, and hormones levels on the domains of AUD.

Forty-eight AUD patients [male (n=34) and female (n=14)] aged 23-63 yrs. were grouped categorically using the Clinical Institute Withdrawal Assessment of alcohol scale (CIWA) as either clinically significant CIWA group (CS-CIWA [score>10] Gr.1 [n=22]), and clinically not-significant group (NCS-CIWA [score≤10] Gr.2 [n=26]). A sub-set of 13 AUD patient were also tested for reward response for drug-seeking using Penn-Alcohol Craving Score (PACS). Clinical data and blood samples were collected upon enrollment. Blood samples were analyzed for pro-inflammatory cytokines, and hormones, and markers of intestinal permeability. CIWA, 90-day timeline followback (TLFB90), and lifetime drinking history (LTDH) were also collected for comparison.

As expected, recent and chronic heavy drinking were significantly higher in Gr.1: HDD90 (heavy drinking days), NDD90 (number of drinking days), as was LTDH, especially in Gr.1 females. Further, in Gr.1, adiponectin (associated with withdrawal) was significantly higher; and numerically higher levels of lipopolysaccharide (LPS) and LPS-binding protein (LBP) were also reported. Gr.1 patients exhibited higher effects of association on the withdrawal-associated depression domain for the parameters of LPS, sCD14, IL-6 and IL-8. Leptin also showed a significantly high effect of association with HDD90 in those AUD patients with craving. The craving domain (assessed by PACS, Penn-Alcohol Craving Scale) could be described as a gut-immune-brain model by the gut-dysregulation (LBP and Leptin) markers, and specific pro-inflammatory activity (IL-1β and TNF-α). Such pathway model describes the heavy drinking phenotype, HDD90 with even higher effects (R^2^=0.955, p=0.006) in the AUD patients who had higher ratings for craving (PACS>5).

Interaction of gut-dysfunction, cytokines involved in both inflammation and in mediating-chemotactic activity constitute a novel pathophysiological gut-brain axis for withdrawal, and alcohol-associated depression and craving domains of AUD. AUD patient with higher craving show higher reinforcing effects of the gut-brain axis response for heavy drinking.

## Introduction

Alcohol use disorder (AUD) is a mental health condition that is characterized by heavy and chronic drinking ^1^. Several pathological mechanisms could be associated uniquely with the reward, ^2^ reinforcement, ^3^ craving ^4^ and withdrawal ^5^ and other domains of AUD. Most treatment/mechanistic studies have targeted AUD pathophysiology based on the influence of alcohol consumption on the brain function measures ^6^. Newer studies reported altered gut-permeability and gut barrier dysfunction, and increased circulating lipopolysaccharides (LPS, gut-derived bacterial products) due to chronic alcohol consumption, and the potentially proximal association of such markers with the behavioral presentation of AUD ^7,8^.

Multiple and highly complex gut-brain axis pathways involve brain biochemistry and neuro-inflammatory mediators ^9^. We recently reported altered pro-inflammatory activity and gut-barrier function in heavy drinkers ^10^. Excessive alcohol consumption causes altered gut-dysbiosis,^11^ intestinal permeability, and gut-derived inflammation ^7,12,13^ that could be related to the chronic and heavy drinking patterns ^10,14^. These changes result in translocation of gut/bacterial derived inflammatory mediators, such as endotoxin (LPS), resulting in gut-derived inflammation ^15^. This has been postulated to be involved in the neuroinflammation of AUD ^16^, though mechanism of gut-barrier dysfunction and proinflammatory response likely constituting a gut-brain axis in AUD is unclear. Thus, understanding the gut-brain axis in the pathology of AUD and its therapy is important.

Chronic and heavy drinking, ^17–19^ as well as frequent episodes of relapse ^20–22^ observed in AUD, limit the efficacy of treatment ^23,24^. Alcohol withdrawal is an essential feature of AUD that could predict the rate and frequency of relapse ^25^. Symptoms of depression are occasionally reported during alcohol withdrawal ^11^; and the explanation of withdrawal associated depression could be rooted to the uniqueness in the neuroinflammatory response ^26^ found in AUD. The role of craving on alcohol relapse ^21^ and involvement of the potential neuro-circuitry ^27^ has been reported recently. Craving for alcohol along with alcohol withdrawal is a well-documented risk factor for subsequent relapse in alcohol use disorder ^28^. Notably, craving is a standalone symptom of AUD as well ^29^ that could manifest phenotypically as the exogenous reward associated presentation of AUD ^14^. Thus, there are reported evidences that indicate towards the role of gut-brain axis and pro-inflammatory activity involved with the AUD domains such as withdrawal, depression and craving in a well-described cohort has largely remained understudied so far ^30^.

The primary aim of this study was to identify the role of gut-brain axis and the pro-inflammatory activity on the domains of alcohol withdrawal, withdrawal associated depression and craving, and craving independently observed in the AUD patients. We also identified and characterized potential blood biomarkers of the gut-barrier dysfunction, endotoxemia, and inflammation involved in the gut-brain axis of AUD. Lastly, we evaluated the involvement of sex, age, or other demographics and drinking patterns in the gut-brain axis in patients with AUD.

## Patients and Methods

### Patient Recruitment

This study was approved by the Institutional Review Board (IRB) of the University of Louisville, Louisville, KY USA, and the Central Neuroscience (CNS) IRB of the National Institute on Alcohol Abuse and Alcoholism, Bethesda MD USA. The study has been indexed at the National Clinical Trial website (www.clinicaltrials.gov): NCT00106106. All AUD patients consented to participate in the study before the collection of clinical and research data, and bodily samples. Forty-eight subjects with AUD, male (n=34) and female (n=14), aged 23 – 63 yrs. participated in this study. Subjects were diagnosed with alcohol use disorder according to DSM-IV, based on the alcohol dependence module of the SCID I-interview, and alcohol withdrawal for either of the two eligibility criteria: (1) clinically manifest significant alcohol withdrawal symptoms, with or without detectable blood alcohol concentrations (BACs); or (2) in the absence of the above, current intoxication above 0.1 g/dl BAC, self-reported history of continuous alcohol intake for more than the past one month, as well as self-reported previous episodes of significantly distressful alcohol withdrawal symptoms. More information on patient participation and enrollment can be obtained in previous publications ^17,31,32^.

*Rest of the Methods is available in the Supplementary Section*.

## Results

### Demographics, Drinking, and Nutritional Assessment

Demographic measures (Age and BMI) were similar in both the study groups. CIWA score in Gr.1 (with Clinically Significant [CS] CIWA scores) patients (45.34% of the overall study participants) was three-fold higher than in Gr.2 (Clinically Not Significant [NCS]) patients (Table 1), as expected. Also, as would be likely, all heavy drinking markers were numerically higher in Gr.1. Notably among these, the HDD90 and NDD90 (derived from the TLFB90 assessment), and LTDH (longitudinal/chronic drinking) drinking markers were significantly higher in Gr.1 (Table 1). LTDH was also significantly higher (p=0.050) in males compared to the females of the Gr.1. Gr.1 females showed more severe patterns of alcohol drinking than the males of Gr.1 (Table 1). We did not find any other sex-differences in drinking markers in Gr.1.

**Table 1.**
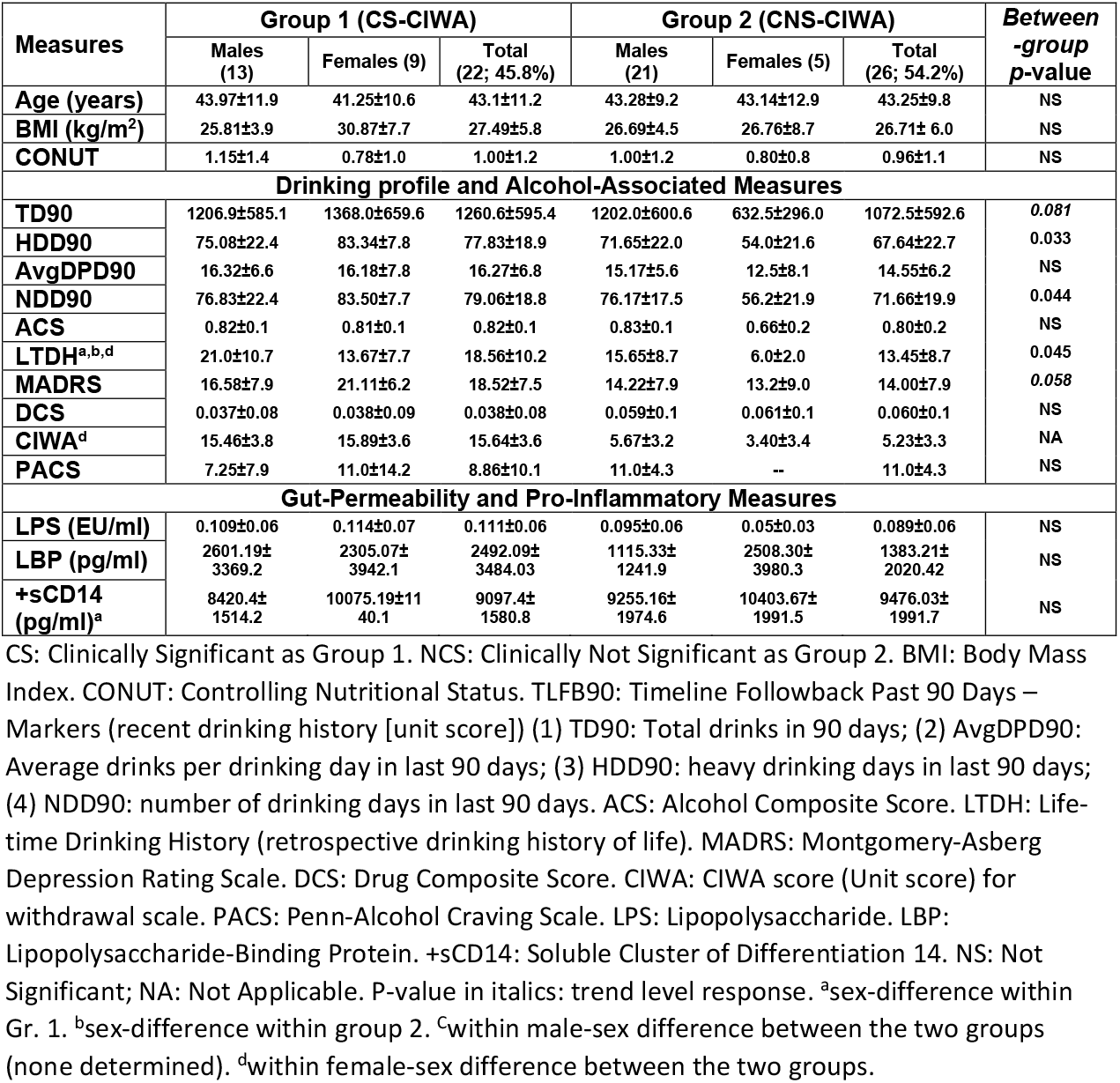
Demographics, markers of recent and long-term drinking history, withdrawal score, and gut permeability measures in AUD patients grouped by the presence of clinically relevant withdrawal. AUD patients are categorized by sex within each of the primary groups.

Gr.1 females drank more than double in the past 90 days, as well as more than double the number of years than the Gr.2 females (Table 1). Gr.1 females also had 37.5% higher Montgomery-Asberg Depression Rating Scale (MADRS) scores, and ~5-fold higher CIWA scores than the Gr.2 females. Gr.1 males exhibited ~3-fold elevated CIWA scores than the Gr.2 males. On the other hand, Gr.2 males drank almost double the amount that the females in Gr.2 drank in the past 90 days (TD90 showed trend toward statistical significance, p=0.052); and NDD90 was also higher in Gr.2 males vs. females (trend level, p=0.056). LTDH and Alcohol Composite Score (ACS) were significantly higher in the males of Gr.2 compared to their female counterparts, as well (Table 1). There were no differences in the CONUT scores between the two groups, or sex-differences within each group.

### Withdrawal Markers, Proinflammatory Activity, and Gut-Dysfunction Response

Adiponectin, a biomarker for alcohol withdrawal,^33^ was significantly (p=0.013) elevated in Gr.1, and the increase was also approximately 2-fold more compared to Gr.2 (Fig. 1a). Gr.1 females exhibited ~37% higher levels of adiponectin than the Gr.1 males (Table 1). Importantly, Gr.1 females also had more than 2-fold higher adiponectin levels than the Gr.2 females (Table 1), this difference was not as well-defined in the males.

**Figure legend 1.**
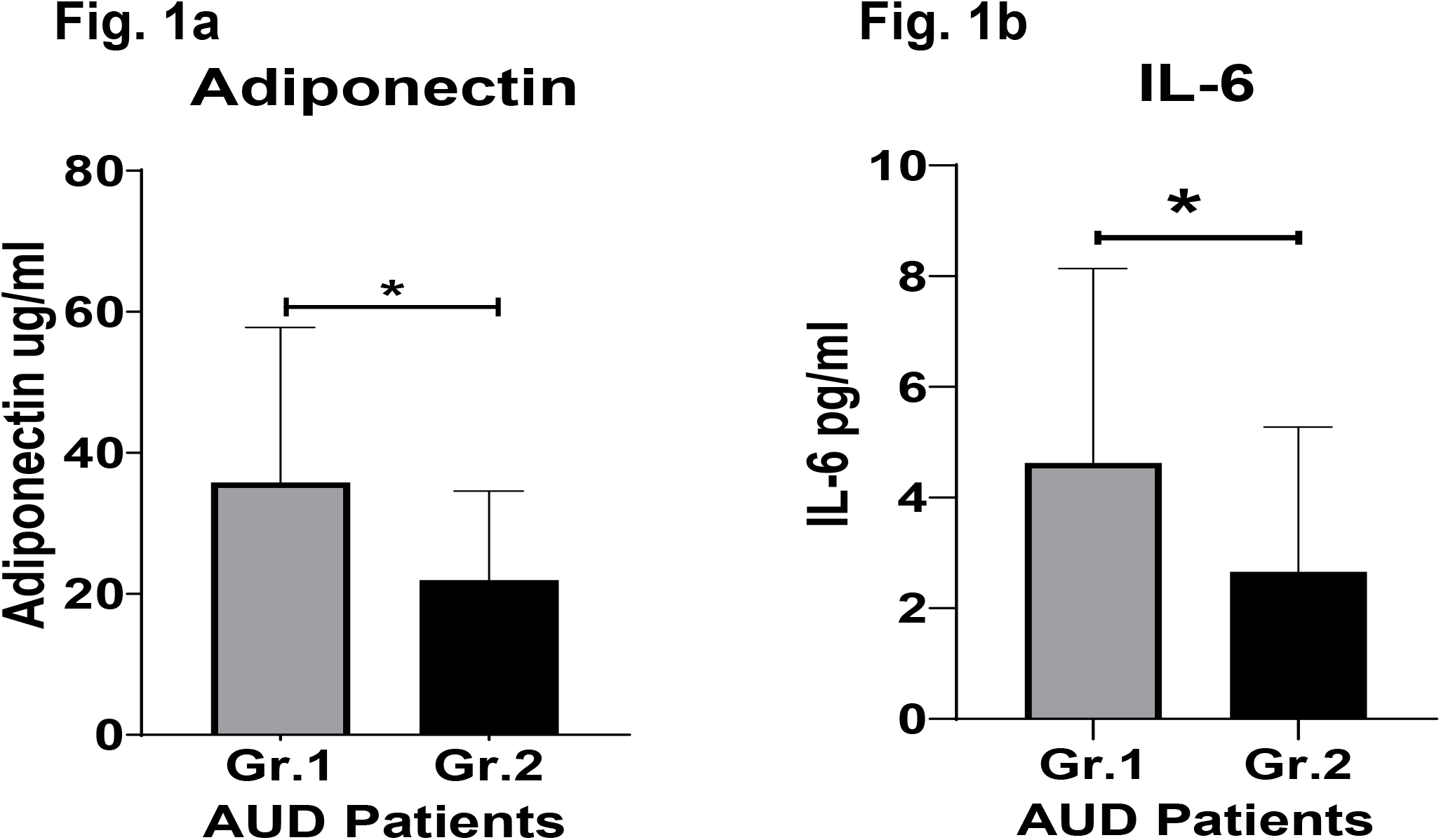
Proinflammatory cytokine and protein hormone activity in AUD patients grouped by clinically significant (CS) level of alcohol withdrawal identified using CIWA scores. Fig. 1a: Adiponectin levels. Fig. 1b: IL-6 levels. Data presented as M±SD. Statistical significance was set at p < 0.05. ** p<0.01. * p<0.05.

Gr.1 had a 2-fold higher IL-6 levels compared to Gr.2 (Fig. 1b). Notably, in Gr.1, the females exhibited ~57% higher levels of IL-6 than their male counterparts (Table 1). Gr.1 females showed numerically higher IL-8 levels (trend level of significance, p=*0*.*070*) than the males in Gr.1 (Table 2); however, due to huge variability, it did not reach statistical significance.

**Table 2.** Candidate pro-inflammatory cytokines, and hormones in AUD patients grouped by the presence of clinically relevant withdrawal. AUD patients are divided by sex within each of the primary groups.

| Measures | Group 1 (CS-CIWA) |  |  | Group 2 (CNS-CIWA) |  |  | *Between-group p-value* |
| --- | --- | --- | --- | --- | --- | --- | --- |
|  | Males (13) | Females (9) | Total (22; 45.8%) | Males (21) | Females (5) | Total (26; 54.2%) |  |
| IL-6 (pg/ml) | 3.77±2.2 | 5.92±4.8 | 4.62±3.5 | 2.70±2.7 | 2.52±2.7 | 2.66±2.6 | p=0.039 |
| TNF-α (pg/ml) | 1.80±0.8 | 2.02±0.9 | 1.89±0.8 | 1.90±0.6 | 1.95±2.0 | 1.91±1.0 | NS |
| IL-1β (pg/ml) | 0.52±0.3 | 0.48±0.6 | 0.51±0.4 | 0.52±0.3 | 0.56±0.4 | 0.52±0.3 | NS |
| MCP-1 (pg/ml)<sup>a</sup> | 110.03±59.9 | 98.37±39.1 | 105.37±51.7 | 100.14±31.4 | 182.3±121.4 | 117.3±67.0 | NS |
| IL-8 (pg/ml) | 4.49±2.6 | 20.16±28.4 | 10.76±19.1 | 4.28±3.3 | 5.96±7.2 | 4.61±4.2 | NS |
| Leptin (pg/ml) | 3074.5±2631.7 | 7179.5±4651.3 | 4716.5±4029.5 | 5766.5±6419.2 | 10125.2±10752.4 | 6674.5±7458.2 | NS |
| Adiponectin (μg/ml) | 30.95±19.1 | 42.5±25.2 | 35.79±22.0 | 22.36±12.5 | 20.32±14.6 | 21.93±12.6 | p=0.013 |

CS: Clinically Significant withdrawal score as Group 1 (Gr.1). NCS: Clinically Not Significant withdrawal score as Group 2 (Gr.2). CIWA: Clinical Institute Withdrawal Assessment Alcohol Scale. NS: Not Significant; NA: Not Applicable. P-value in italics: trend level response. <sup>a</sup>sex-difference within group 2.

Gut permeability markers (especially LBP) were numerically higher in Gr.1 (albeit did not reach statistical significance likely due to larger standard deviation in both the groups, Table 1). LBP was ~2.3 fold higher in Gr.1 males compared to the males of Gr.2 (trend level, p=0.077). Gr.1 females showed significantly higher +sCD14 levels than their Gr.1 male counterparts (Table 1). LBP was significantly associated (p=0.022) with heavy drinking pattern, HDD90 only in Gr.1 (R^2^=0.351, p=0.010) (data not shown).

HDD90 and TNF-α values showed statistically significant, albeit with mild effects across all the study patients (Fig. 2a). However, the underlying effect of this association had a significance between HDD90 and TNF-α (R^2^=0.207 at p=0.050) in the Gr.1 group (Fig. 2b), though this was not significant in Gr.2 (Data not shown).

**Figure legend 2.**
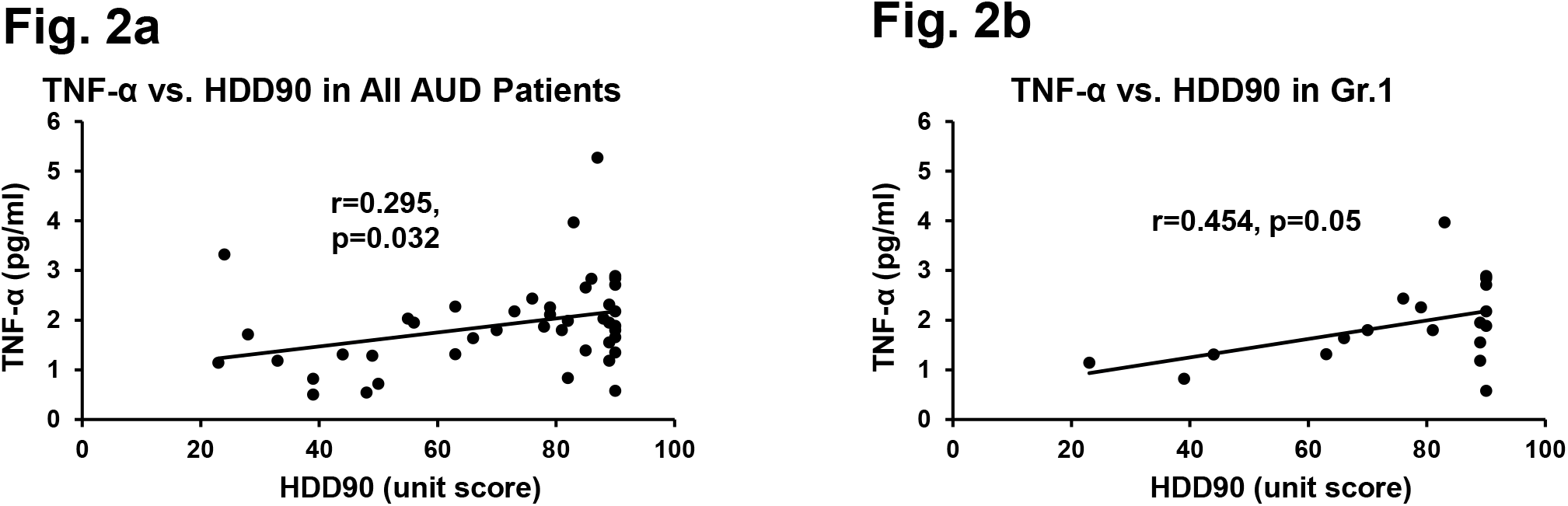
Association of heavy drinking and pro-inflammatory activity. Fig. 2a: TNF-α and HDD90 in all AUD patients. Fig. 2b: TNF-α and HDD90 in Gr. 1 AUD patients with clinically relevant withdrawal scores. r denotes the correlation coefficient of relation. Statistical significance was set at p < 0.05.

### Alcohol Withdrawal, Drinking Patterns and Gut-Brain Axis Markers

Withdrawal score CIWA and adiponectin were significantly associated with the chronic drinking marker, LTDH, in all patients with AUD (Fig. 3a and Fig. 3b respectively). CIWA showed significant association with adiponectin (p=0.003) in all AUD patients (Fig. 3c) as well.

**Figure legend 3.**
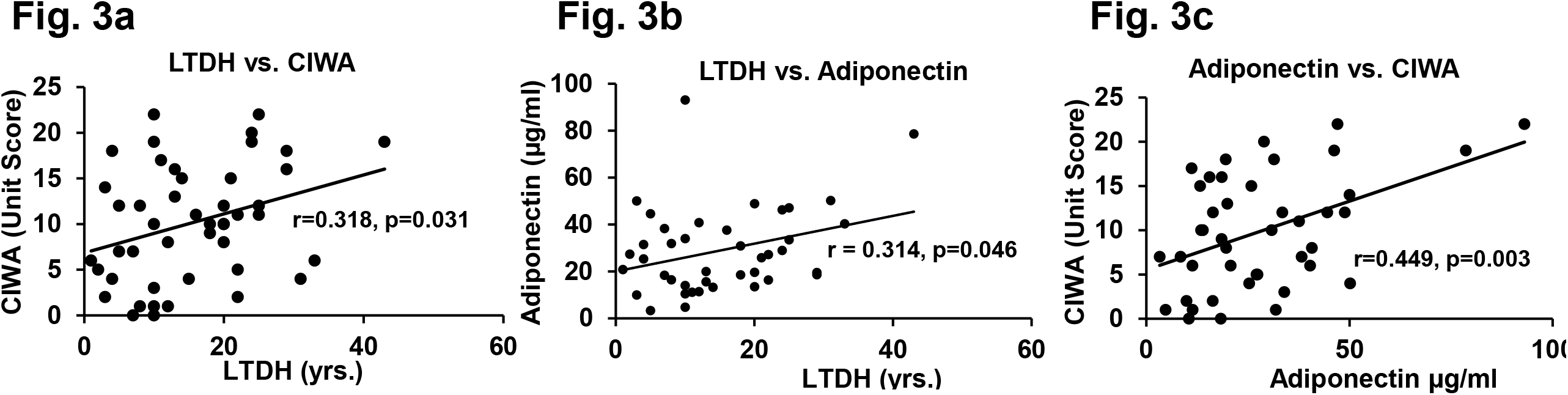
Association of withdrawal score with chronic drinking measures, level of adiponectin in all the AUD patients. Fig. 3a: Association of chronic drinking (LTDH) and CIWA. Fig. 3b: Association of chronic drinking (LTDH) and Adiponectin. Fig. 3c: Association of CIWA and plasma adiponectin level (A laboratory marker of withdrawal). Statistical significance was set at p < 0.05.

Only the Gr.1 (CS) patients showed a stepwise increase in the effects of association of CIWA scores and adiponectin with LPS (R^2^=0.474, p=0.006); and this association effect was augmented further with respect to pro-inflammatory activity in a stepwise manner for IL-6 (R^2^=0.494, p=0.015) and IL-8 (R^2^=0.510, p=0.031) (Fig. 4a, schematic). Gr.2 patients did not exhibit such arrangement of gut-immune-brain response (Fig. 4b).

**Figure legend 4.**
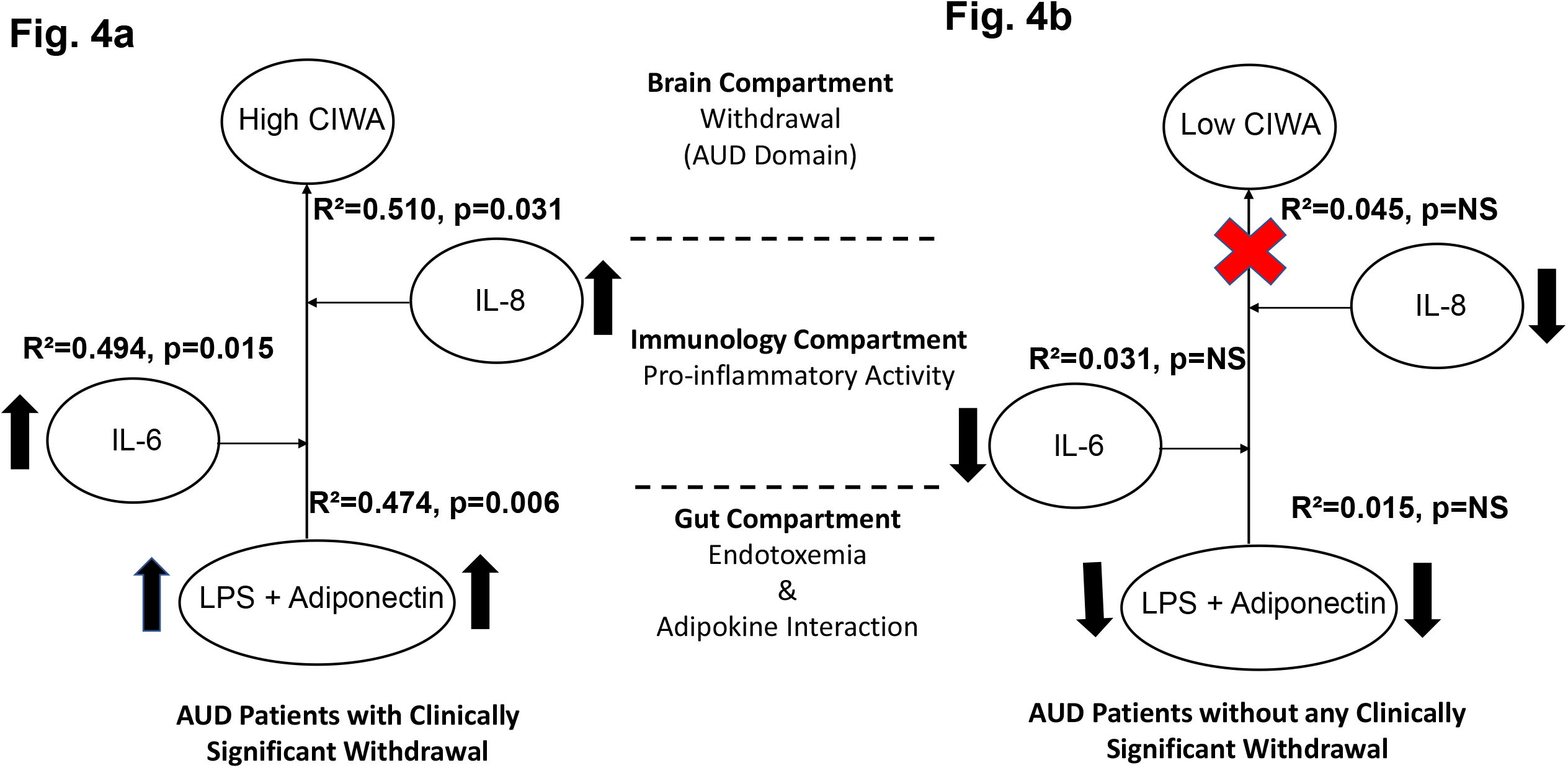
Model for gut-brain axis illustrating effects and schema of gut-dysfunction, proinflammatory and adiponectin activity on alcohol withdrawal in (Fig. 4a) AUD patients with clinically significant CIWA scores; and (Fig. 4b) without clinically significant withdrawal. Statistical significance was set at p < 0.05. Arrows indicate higher levels of specific measures observed in Gr. 1 than the Gr. 2. Upward arrows show comparative increase between the two sub-figures. Red cross sign denotes no effect. Effect size ranges: mild <0.2; 0.2<moderate>0.7; robust>0.7.

### Depression Associated with Alcohol Withdrawal, Drinking Patterns and Gut-Brain Axis Markers

MADRS score was significantly associated with +sCD14 response (a gut-dysfunction measure that facilitates cellular responses to LPS) with mild effects (R^2^= 0.208 at p=0.038) in Gr. 1 that augmented (R^2^= 0.217 at p=0.043) in conjunction with the confounder LPS (circulating LPS indicates gut barrier dysfunction/leakiness). Ultimately, this arrangement of the gut-dysfunction on MADRS showed much higher effects in the context of candidate proinflammatory activity in a stepwise increasing fashion, with IL-6 (R^2^=0.546, p=0.007) and IL-8 (R^2^=0.563, p=0.015) respectively (Fig. 5a). Such strong gut-brain axis signature on depression domain was not present in Gr. 2 (Fig. 5b).

**Figure legend 5.**
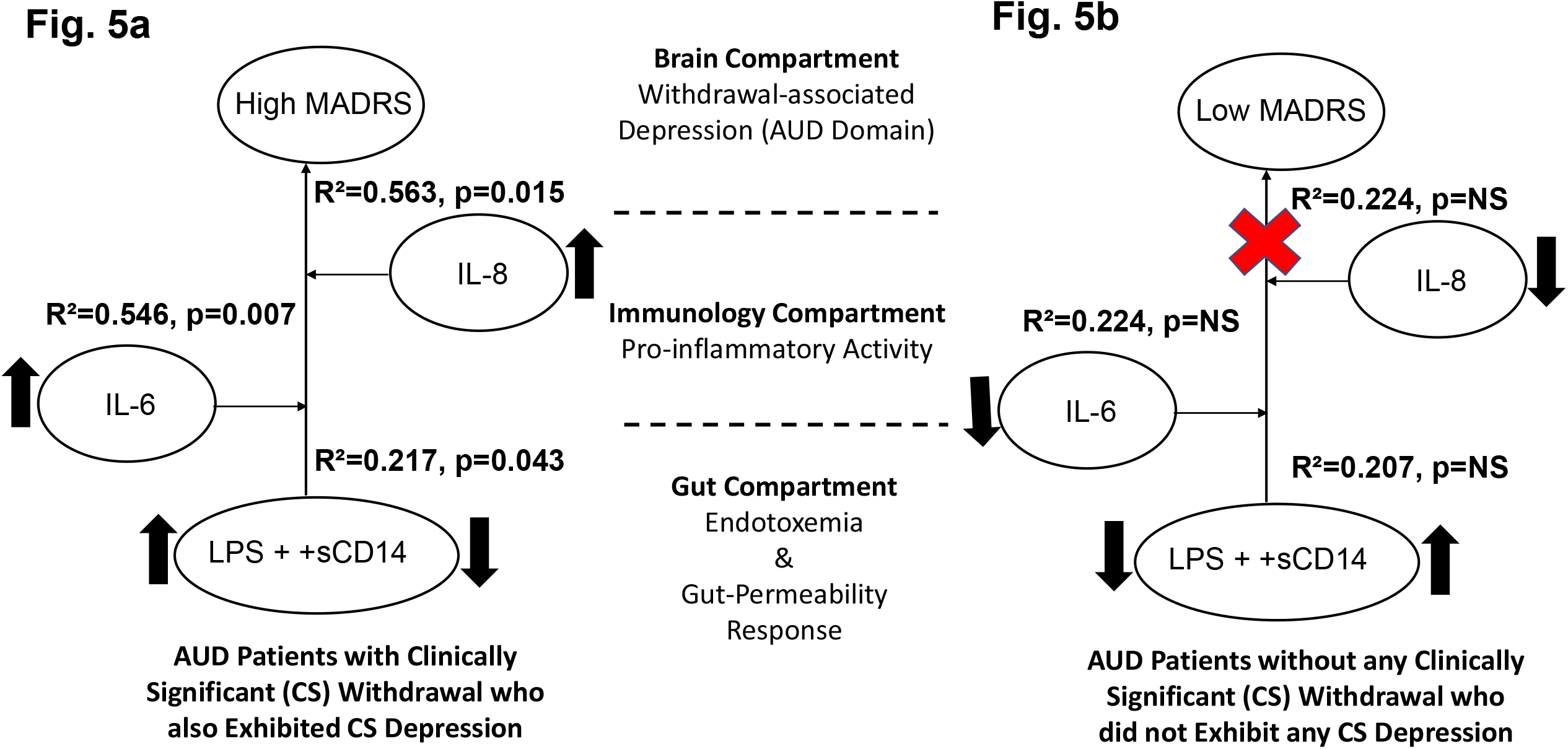
Model for gut-brain axis involving gut-dysfunction, proinflammatory activity on MADRS scores in (Fig. 5a) AUD patients with clinically significant withdrawal (Gr.1); and (Fig. 5b) AUD patients without clinically significant withdrawal (Gr. 2). Statistical significance was set at p < 0.05. Upwards arrows indicate higher levels of specific measures observed in Gr. 1 compared with the Gr. 2 values. Red cross sign denotes no effect. Effect size ranges: mild <0.2; 0.2<moderate>0.7; robust>0.7.

### ICraving & Leptin Response, Drinking Patterns and Gut-Brain Axis Markers

Gr.1 leptin levels (a marker of craving) were lower, around 2/3 times that of Gr.2 levels (Table 2). An inverse relationship between LTDH and leptin levels were significant (R^2^=0.201, p=0.047) (Fig. 6a) only in Gr. 1. In the Gr. 2 AUD patients without any withdrawal, higher levels of leptin and lower HDD90 values showed significant association, p=0.038 at mild effects, R^2^=0.14 (data not shown); such response was not significant in Gr.1. Notably, Leptin and HDD90 showed a significant inverse albeit mild association exclusively among the males of this study (regardless of withdrawal), R^2^=0.181 at p=0.017 (Fig. 6b). Leptin levels also showed sexual dimorphism, and corresponding vulnerability towards males ^34^ of the AUD group with clinically significant withdrawal. These values were lower than half that of the females of the same group, at p=0.021 (Table 2).

**Figure legend 6.**
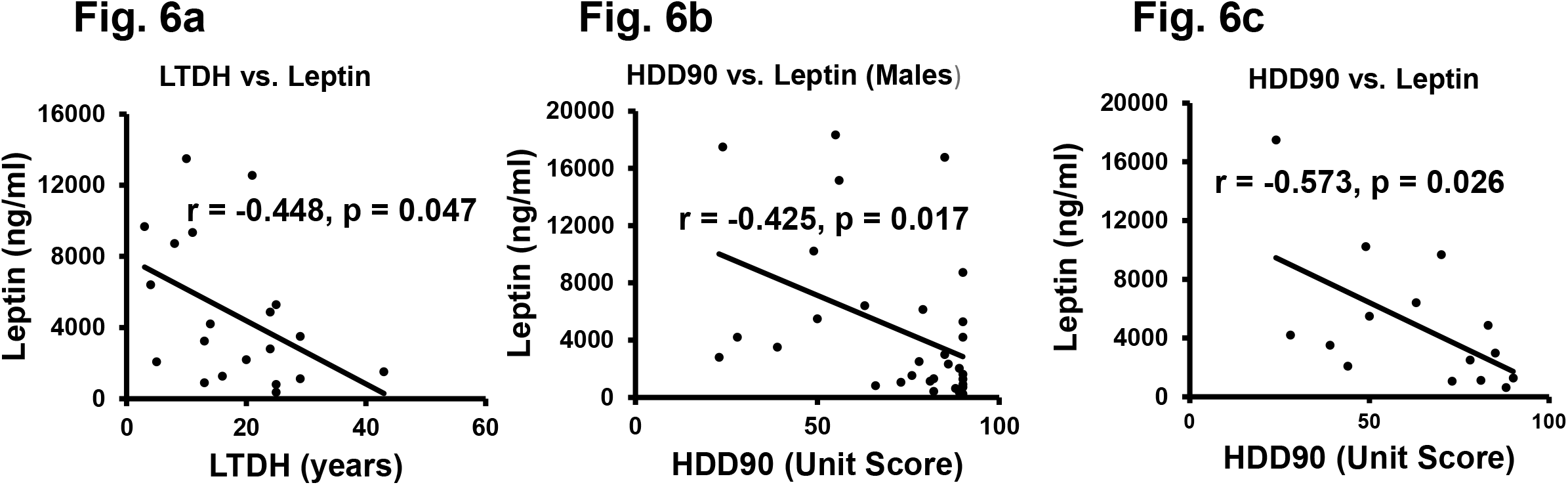
Association of patterns of alcohol intake and leptin in AUD patients. Fig. 6a. Lifetime drinking history (LTDH), chronic drinking markers showed negative association with Leptin in Gr.1 AUD patients exhibiting withdrawal. Fig. 6b: Recent heavy drinking pattern in the past 90 days (HDD90) showed negative relationship with Leptin in all the male AUD patients of this study. Fig.6c: HDD90 and Leptin showed higher effect of negative association among the AUD patients who reported on the PACS score. Statistical significance was set at p < 0.05.

Sub-set analyses of 16 AUD patients tested for craving showed some unique and significant gut-brain response and involvement of pro-inflammatory cytokines when assessed by the PACS scores. There were 13 males and three females, who did not show much numerical or statistical sex-difference in PACS scores. AUD patients who had craving recorded using PACS scores showed a significant inverse association between leptin and HDD90, R^2^=0.328 at p=0.026 (Fig. 6c). PACS was significantly and positively associated p=0.011 (R^2^=0.40) with the gut-permeability marker, LBP as well (data not shown).

We evaluated the gut-immune-brain connection in the context of craving and identified the role of LBP and leptin (gut response) along with TNF-α and IL-1β (immunological status) contributing with very high effects and significantly predicting the PACS score (Fig. 7a). When we compared the same gut-brain interaction to evaluate the drinking response of craving, HDD90, we found an even higher effect size (Fig. 7b) in AUD patients with PACS>5, this association was not observed in patients with 5<PACS score.

**Figure legend 7.**
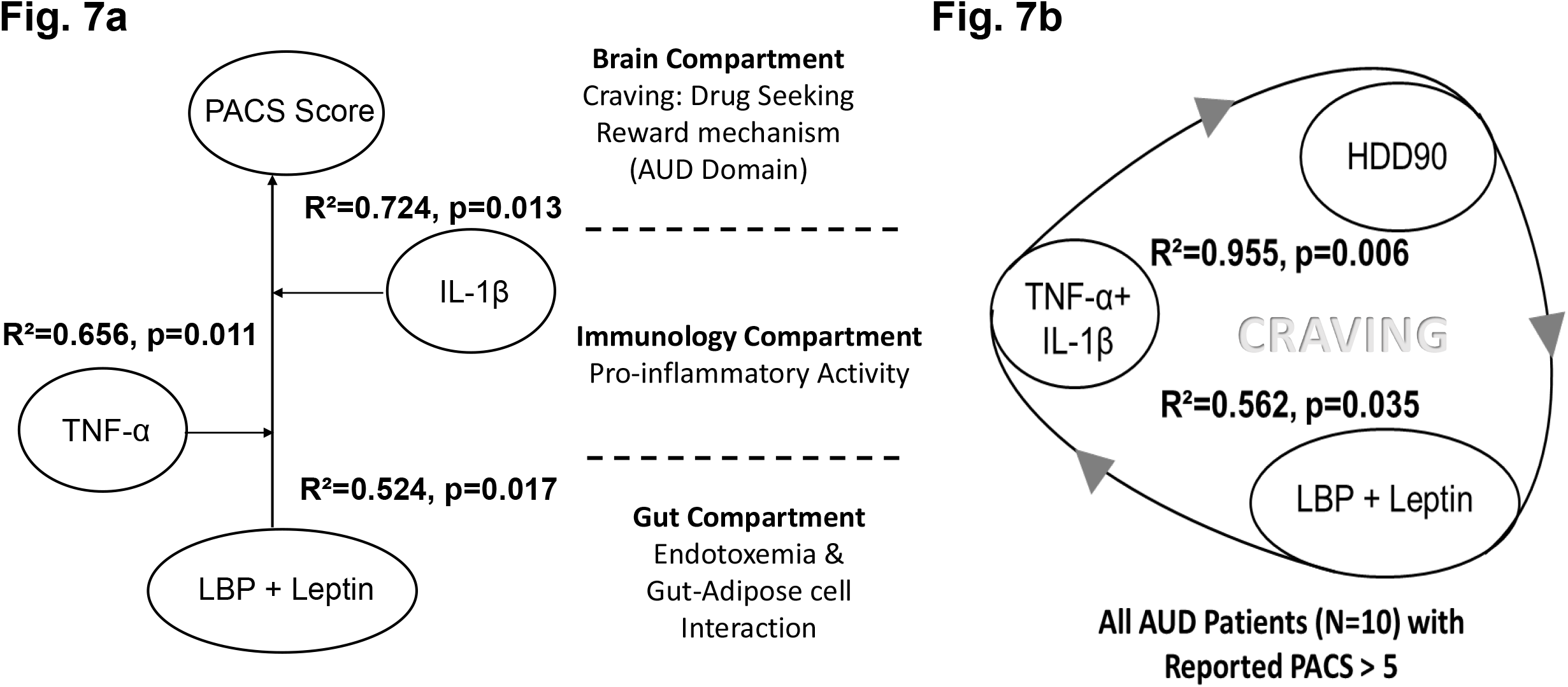
Fig. 7a: Model for gut-brain axis involving gut-dysfunction, proinflammatory activity on the craving scale (PACS) in a sub-set of AUD patients of this study. Fig. 7b: Corresponding model for heavy drinking days (HDD90) marker of heavy drinking (as an exophenotype determinant) exhibited very high effects in AUD patients who exhibited PACS>5. Statistical significance was set at p < 0.05. Effect size ranges: mild <0.2; 0.2<moderate>0.7; robust>0.7.

## Discussion

Our study revealed that AUD patients with clinically significant CIWA scores presented with exacerbated heavy and chronic drinking patterns at the time of intake. Drinking profile of the study patients exhibited uniquely high heavy drinking patterns, especially HDD90,NDD90 and TD90. This suggests recent noteworthy changes in drinking behavior occurring past 90 days of assessment. The uniqueness in the combination of chronicity of drinking coupled with recent acute heavy drinking pattern could play an important role in characterizing various facets of AUD including specific withdrawal symptoms, depression, craving. We found that the female AUD patients with clinically significant withdrawal symptoms drank more heavily than their male counterparts (with similar withdrawal scores), suggesting that they are drinking at levels that might go unnoticed. Importantly, the ongoing adverse influence on other domains of AUD and alcohol-associated organ injury may well be progressing simultaneously either independently or in a comorbid form. Evidently, we found that this group of female patients also showed correspondingly higher MADRS score (depression symptoms). These findings may be related to sex-specific vulnerability of female patients when compared with male counter parts^19,35,36^.

Patients with AUD exhibiting withdrawal exhibited a close association between proinflammatory cytokines and adiponectin. The interaction of adiponectin and LPS, and higher proinflammatory cytokine levels supports the role of proinflammatory activity in withdrawal and supports a possible pathogenic role of gut-brain axis in exacerbating withdrawal. Female patients of our study with high withdrawal showed greater predisposition ^35^ by the changes in gut permeability and proinflammatory cytokine response.

Earlier mechanistic studies delineated AUD pathophysiology targeting the deleterious effects of alcohol on certain brain functions ^6,37^. Later studies have clearly shown that excessive alcohol consumption leads to both increased circulating levels of LPS deriving from gut bacterial products and increased gut permeability ^38^. Subsequent dysregulation in gut-permeability and function that we addressed in context of LPS, and adiponectin could follow-up with the establishment of the pro-inflammatory status. A subsequent alteration in immune response with alcohol consumption leads to pro-inflammatory status ^39^. Several biomarkers were detected at gut, that corresponded well with the cytokine response among the exogenous phenotypic presentation of the candidate domains of AUD. Hillmer et al., showed that acute alcohol consumption alters the levels of peripheral cytokines such as IL-8 and TNF-α ^40^. TNF-α response is an early cell-signaling proinflammatory factor that has been reported at lower levels ^41^ or without significant different levels ^42^ in neurological conditions such as depression. In our study, TNF-α (which was not significantly different from AUD patients without withdrawal ^43^, Table 2) and HDD90 were associated in AUD patients exhibiting withdrawal. This supports the unique characterization of proinflammatory activity with a heavy drinking marker indicating a relationship with alcohol-associated neuroinflammation ^44,45^, leading to or contributing in withdrawal ^11^. We could identify a three compartment response system that could be involved in characterizing the gut-brain axis for any of the domains that we studied. Our findings also highlight the elevated levels of IL-6 and IL-8 that is observed with alcohol intake ^46^. Overproduction of IL-6 (which passes through blood brain barrier ^47^) and IL-8 (which increases the blood-brain barrier permeability ^48^) proceed the initial TNF-α signaling response in the patients with AUD, which might not be as contributing in AUD patients without withdrawal. This could be a major neurobiological key or gateway of withdrawal ^49^ and other domains of AUD ^50^ such as withdrawal associated depression ^51^. However, how these cytokines may manifest with the domain/s of AUD could be a turning point; and thus, becomes a highly important area of investigation. Some of these correlations of cytokine levels and arrangement of gut permeability factors coincide with our earlier observations with hyperhomocysteinemia, gut dysfunctions and markers of excessive drinking ^32^. These intestinal changes as reflected by the alterations in blood markers, also lead to behavioral changes that could correspondingly be observed with the severity of alcohol abuse ^7,8,52^. In our study, we found a derangement of gut-permeability factors and role of proinflammatory cytokines that contributed uniquely as gut-brain axis of AUD in characterizing withdrawal and withdrawal-associated depression in a domain-specific manner.

Under normal circumstances, the human gut maintains its microbiome ^53^. Functional changes of this commensal microbiota lead to several pathogenetic conditions. We identified a corresponding increasing the level of withdrawal with lifetime drinking. Several findings suggest that changes in the gut microbiome (dysbiosis) largely contribute to certain deleterious effects of chronic excessive alcohol intake ^44,54^, and attenuating these changes is a therapeutic option that has great interest ^55^. These reports revealed an important biological association between gut-dysregulation and alcohol intake, and this emphasizes the need for therapeutic intervention on gut microbial targets to treat AUD.

Craving associated with alcohol withdrawal is a well-documented risk factor for subsequent relapse in AUD ^28^. One promising marker of craving, leptin was low in our patients with AUD who exhibited clinical levels of withdrawal. Leptin is an appetite-regulating hormone produced by adipose, brain and gut cells and is known for its association with craving ^56^. Leptin can be synthesized by the stomach and brain ^57,58^, and peripheral leptin could also reach central nervous system via cellular transportation of the blood-brain barrier ^59^. Our findings showed a negative relationship between the markers of alcohol intake and leptin, which has also been reported in one previous study ^60^. Studies have shown that alcohol administration lowered the leptin levels in alcohol-preferring rat and mice model experiments ^61,62^. This could be relatable to our patient population with a heavy and chronic drinking profile. This lowering has been observed in human studies ^63^ previously. One study supported the diurnal and nocturnal effects of alcohol in leptin concentrations ^64^, while another suggested that its availability is higher in cerebrospinal fluid ^61^. We noticed that the females had higher levels of leptin than the males in patients with AUD who regardless of the presence/absence of withdrawal. This is consistent with recent findings on sex-difference with alcohol drinking behavior ^65,66^. None-the-less, it seems that lowering of leptin was more reflective of heavy drinking in males with AUD (regardless of presence/absence of withdrawal) in our study patients.

Craving is an important domain of AUD that may occur independent of withdrawal as well ^36^. It may occur at non-clinical stages even in social drinkers ^67^, but how it differs in manifestation when AUD is diagnosed may be explained by the modifying factors that come into play subsequently. Thus, investigating the role of gut-brain axis specific for craving assessments provided unique reward-associated findings. Our findings had specific commonality for precursor gut-permeability factors and pro-inflammatory cytokine response with other domains. However, subsequent changes in specific metabolites likely contribute to the development of the specific domains. Gut-dysfunction that is marked by LBP response coupled with leptin (likely originating from the alcohol-associated gut-dysfunction) could develop craving with the ongoing dysregulation in the pro-inflammatory cytokine activity. The pro-inflammatory activity that might be involved in craving could have the origin from the dysregulated cytokine response of both TNF-α and IL-1β, uniquely adhered to AUD patients with high craving. As discussed earlier, TNF-α could be involved in the triggering the gut-dysfunction associated immunological dysregulation ^49^ and that perpetuates further with the involvement of IL-1β ^68^ to uniquely identify the immune signaling response of craving in AUD patients. Craving as described by PACS score in our study also showed a gut-brain response with the involvement of immune activity. An exogenous phenotype character of craving was highly reproducible with the heavy drinking marker, HDD90 with the same gut-brain axis model. However, most concerning explanation of such a strong response could present with a vicious cycle of the heavy drinking pattern that may be self-feeding to exacerbate through the gut-brain axis.

The generation of gut microbiota derived neurotransmitters that invoke potent biological effects was demonstrated by several groups ^54,69–72^. We also showed these effects on serotonin pathways previously ^73,74^. Others have examined GABA ^75,76^, dopamine synthesis ^14^ and the release of short chain fatty acids ^77–80^ (derived from the gut microbiome). All these end products have potential neuroactive properties ^55^. Although we have not estimated the levels of these chemicals in the present study, it is evident from our earlier published studies, as well as those by others, that these molecules exert effects on several brain functions especially linked to behavioral alterations^12,18,72^. It is interesting to note that studies in mice have unequivocally shown the critical role of gut microbial catecholamines, which are biologically active, both in the gut as well as in distant brain segments ^81^. As early as 2011, researchers showed that intestinal microbiota critically affect central levels of brain-derived neurotropic factor(s) and behavior in mice ^82^.

There are some limitations in this study. First, as this was a proof-of-principle study, it had a relatively small sample size. Consequentially, many underlying effects are not addressed in this study and should be the scope of future investigations. Group 2 patients with AUD, but without withdrawal symptoms, had only a small sub-group of females; thus, the interpretations for females are limited. However, the focus was on the patients with AUD and withdrawal; for which the study included sufficient number of patients of both sexes. Many studies have shown significant elevation of specific cytokines, such as TNF-α with AUD; however, the current findings support slight lowering, which we have justified with the available literature that also suggest lowering of TNF-α. There is a third direction on it as well, which is that with neuroinflammation, once the blood-brain barrier (BBB) has increased permeability, a lot of cytokines influx into the brain. Thus, there may have been relative lowering of certain cytokines in blood, an investigation that was beyond the scope of this study. A similar limitation could be associated with leptin levels, where a comparative proportion of the leptin may have crossed the BBB in AUD patients with withdrawal. Our study was focused on assessing three domains in AUD patients with clinically significant domain criteria: CIWA scores, depression rating, and craving and severity due to alcohol drinking as the alcohol composite score (please see the supplementary section for results and discussion). Direct assessment of reward, and reinforcement were not within the scope of this study. The identification/role of specific pathogenic gut-microbiota was also not investigated. The latter is an area for future investigation.

Gut-dysfunction due to heavy alcohol drinking, and the gut-brain axis seemed to express the derangement of relevant biomarkers and specific clinical presentation/s in AUD patients for different domains in unique arrangements. In this study, we have begun to characterize the gut-brain axis of alcohol withdrawal, associated depression, and craving developing a gut-brain model of alcohol pathology with three major realms. They are: (1) direct consequence of drinking on withdrawal marker(s), (2) candidate markers of gut dysfunction, and (3) specific cytokine dysregulation; as biological compartments of interest. This study is a proof-of-principle study which shows the need for further in-depth study of this important relation.

## Supporting information

Supplement to GBA

## Data Availability

The datasets analyzed during the current study are available from the corresponding author on reasonable request. Email address for the readers to contact the author to obtain the data.

## Acknowledgments

We thank research/clinical staff of the University of Louisville and National Institutes of Health for their support. We thank Ms. Marion McClain for editorial support for this manuscript.

## Reprint Request

Vatsalya Vatsalya MD PgD MS MSc, Department of Medicine, University of Louisville School of Medicine, 505 S. Hancock St., CTR Room 514A Louisville KY 40202. Landline: 502-852-8928, Fax: 502-852-8927. Cell: 502-488-0446.

## Notes

**Conflicts of Interest:** Over the past 3 years, J.C.V. has acted as a consultant/advisor for KNMP, More Labs, Red Bull, Sen-Jam Pharmaceutical, Toast!, and Tomo. All other authors declare no conflict of interests.

**Project/Grant Support:** Study was supported by NIH: Z99-AA999999, K23AA029198-01 (VV), R15CA170091-01A1 (MK), ZIA AA000466 (VAR), R01AA023190 (WF), P50AA024337-8301 and P20GM113226-6169 (XZ), and P50AA024337, P20GM113226, U01AA026934, U01AA026926, U01AA026980 (CJM), and the VA (CJM). The content is solely the responsibility of the authors and does not necessarily represent the official views of the National Institutes of Health.

### Competing Interest Statement

Over the past 3 years, J.C.V. has acted as a consultant/advisor for KNMP, More Labs, Red Bull, Sen-Jam Pharmaceutical, Toast!, and Tomo. All other authors declare no conflict of interests.

### Funding Statement

Study was supported by NIH: Z99-AA999999, K23AA029198-01 (VV), R15CA170091-01A1 (MK), ZIA AA000466 (VAR), R01AA023190 (WF), P50AA024337-8301 and P20GM113226-6169 (XZ), and P50AA024337, P20GM113226, U01AA026934, U01AA026926, U01AA026980 (CJM), and the VA (CJM). The content is solely the responsibility of the authors and does not necessarily represent the official views of the National Institutes of Health.

### Author Declarations

This study was approved by the Institutional Review Board (IRB) of the University of Louisville, Louisville, KY USA, and the Central Neuroscience (CNS) IRB of the National Institute on Alcohol Abuse and Alcoholism, Bethesda MD USA. The study has been indexed at the National Clinical Trial website (www.clinicaltrials.gov): NCT00106106. All patients included in this study consented to participate before the beginning of the study.

## References

1. Kranzler HR, Soyka M. Diagnosis and pharmacotherapy of alcohol use disorder: a review. Jama. 2018;320(8):815–824.

2. Alba-Ferrara L, Müller-Oehring E, Sullivan E, Pfefferbaum A, Schulte T. Brain responses to emotional salience and reward in alcohol use disorder. Brain imaging and behavior. 2016;10(1):136–146.

3. McBride WJ, Lovinger DM, Machu T, et al. Serotonin-3 receptors in the actions of alcohol, alcohol reinforcement, and alcoholism. Alcoholism: Clinical and Experimental Research. 2004;28(2):257–267.

4. Simon J, Etienne A-M, Bouchard S, Quertemont E. Alcohol craving in heavy and occasional alcohol drinkers after cue exposure in a virtual environment: The role of the sense of presence. Frontiers in Human Neuroscience. 2020;14.

5. Anton RF, Latham P, Voronin K, et al. Efficacy of Gabapentin for the Treatment of Alcohol Use Disorder in Patients With Alcohol Withdrawal Symptoms: A Randomized Clinical Trial. JAMA Internal Medicine. 2020;180(5):728–736.

6. Gilpin NW, Koob GF. Neurobiology of alcohol dependence: focus on motivational mechanisms. Alcohol Research & Health. 2008;31(3):185.

7. Leclercq S, Matamoros S, Cani PD, et al. Intestinal permeability, gut-bacterial dysbiosis, and behavioral markers of alcohol-dependence severity. Proceedings of the National Academy of Sciences. 2014;111(42):E4485–E4493.

8. Leclercq S, De Saeger C, Delzenne N, de Timary P, Stärkel P. Role of inflammatory pathways, blood mononuclear cells, and gut-derived bacterial products in alcohol dependence. Biological psychiatry. 2014;76(9):725–733.

9. O’connor J, Lawson M, Andre C, et al. Lipopolysaccharide-induced depressive-like behavior is mediated by indoleamine 2, 3-dioxygenase activation in mice. Molecular psychiatry. 2009;14(5):511–522.

10. Kirpich IA, McClain CJ, Vatsalya V, et al. Liver injury and endotoxemia in male and female alcohol-dependent individuals admitted to an alcohol treatment program. Alcoholism: Clinical and Experimental Research. 2017;41(4):747–757.

11. Gorky J, Schwaber J. The role of the gut–brain axis in alcohol use disorders. Progress in Neuro-Psychopharmacology and Biological Psychiatry. 2016;65:234–241.

12. Leclercq S, de Timary P, Delzenne NM, Stärkel P. The link between inflammation, bugs, the intestine and the brain in alcohol dependence. Translational psychiatry. 2017;7(2):e1048–e1048.

13. Kelley KW, Dantzer R. Alcoholism and inflammation: neuroimmunology of behavioral and mood disorders. Brain, behavior, and immunity. 2011;25:S13–S20.

14. Vatsalya V, Gowin JL, Schwandt ML, et al. Effects of varenicline on neural correlates of alcohol salience in heavy drinkers. International Journal of Neuropsychopharmacology. 2015;18(12).

15. Bishehsari F, Magno E, Swanson G, et al. Alcohol and gut-derived inflammation. Alcohol research: current reviews. 2017;38(2):163.

16. Mayfield J, Ferguson L, Harris RA. Neuroimmune signaling: a key component of alcohol abuse. Current opinion in neurobiology. 2013;23(4):513–520.

17. Vatsalya V, Kong M, Cave MC, et al. Association of serum zinc with markers of liver injury in very heavy drinking alcohol-dependent patients. The Journal of nutritional biochemistry. 2018;59:49–55.

18. Vatsalya V, Hassan HZ, Kong M, et al. Exacerbation of Hangover Symptomology Significantly Corresponds with Heavy and Chronic Alcohol Drinking: A Pilot Study. Journal of clinical medicine. 2019;8(11):1943.

19. Vatsalya V, Song M, Schwandt ML, et al. Effects of sex, drinking history, and omega-3 and omega-6 fatty acids dysregulation on the onset of liver injury in very heavy drinking alcohol-dependent patients. Alcoholism: Clinical and Experimental Research. 2016;40(10):2085–2093.

20. Becker HC. Alcohol dependence, withdrawal, and relapse. Alcohol Research & Health. 2008.

21. Bottlender M, Soyka M. Impact of craving on alcohol relapse during, and 12 months following, outpatient treatment. Alcohol and Alcoholism. 2004;39(4):357–361.

22. Breese GR, Sinha R, Heilig M. Chronic alcohol neuroadaptation and stress contribute to susceptibility for alcohol craving and relapse. Pharmacology & therapeutics. 2011;129(2):149–171.

23. Marlatt GA, Witkiewitz K. Relapse prevention for alcohol and drug problems. 2005.

24. Streeton C, Whelan G. Naltrexone, a relapse prevention maintenance treatment of alcohol dependence: a meta-analysis of randomized controlled trials. Alcohol and Alcoholism. 2001;36(6):544–552.

25. Loeber S, Duka T, Welzel Márquez H, et al. Effects of repeated withdrawal from alcohol on recovery of cognitive impairment under abstinence and rate of relapse. Alcohol and Alcoholism. 2010;45(6):541–547.

26. Hurley LL, Tizabi Y. Neuroinflammation, neurodegeneration, and depression. Neurotoxicity research. 2013;23(2):131–144.

27. Heinz A, Beck A, Grüsser SM, Grace AA, Wrase J. Identifying the neural circuitry of alcohol craving and relapse vulnerability. Addiction biology. 2009;14(1):108–118.

28. Bottlender M, Soyka M. Impact of craving on alcohol relapse during, and 12 months following, outpatient treatment. Alcohol and alcoholism (Oxford, Oxfordshire). 2004;39(4):357–361.

29. Hartwell EE, Ray LA. Craving as a DSM-5 symptom of alcohol use disorder in non-treatment seekers. Alcohol and Alcoholism. 2018;53(3):235–240.

30. Morris LS, Voon V, Leggio L. Stress, motivation, and the gut–brain axis: a focus on the ghrelin system and alcohol use disorder. Alcoholism: Clinical and Experimental Research. 2018;42(8):1378–1389.

31. Vatsalya V, Gala KS, Mishra M, et al. Lower Serum Magnesium Concentrations are associated With Specific Heavy Drinking Markers, Pro-Inflammatory Response and Early-Stage Alcohol-associated Liver Injury. Alcohol and Alcoholism. 2020;55(2):164–170.

32. Vatsalya V, Gala KS, Hassan AZ, et al. Characterization of Early-Stage Alcoholic Liver Disease with Hyperhomocysteinemia and Gut Dysfunction and Associated Immune Response in Alcohol Use Disorder Patients. Biomedicines. 2021;9(1):7.

33. Buechler C, Schäffler A, Johann M, et al. Elevated adiponectin serum levels in patients with chronic alcohol abuse rapidly decline during alcohol withdrawal. Journal of gastroenterology and hepatology. 2009;24(4):558–563.

34. Montague CT, Prins JB, Sanders L, Digby JE, O’Rahilly S. Depot-and sex-specific differences in human leptin mRNA expression: implications for the control of regional fat distribution. Diabetes. 1997;46(3):342–347.

35. Vatsalya V, Bin Liaquat H, Ghosh K, Prakash Mokshagundam S, J McClain C. A review on the sex differences in organ and system pathology with alcohol drinking. Current drug abuse reviews. 2016;9(2):87–92.

36. Gowin JL, Sloan ME, Stangl BL, Vatsalya V, Ramchandani VA. Vulnerability for alcohol use disorder and rate of alcohol consumption. American Journal of Psychiatry. 2017;174(11):1094–1101.

37. Carvalho AF, Heilig M, Perez A, Probst C, Rehm J. Alcohol use disorders. The Lancet. 2019;394(10200):781–792.

38. Bode C, Bode JC. Effect of alcohol consumption on the gut. Best practice & research Clinical gastroenterology. 2003;17(4):575–592.

39. Bjørkhaug ST, Neupane SP, Bramness JG, et al. Plasma cytokine levels in patients with chronic alcohol overconsumption: relations to gut microbiota markers and clinical correlates. Alcohol. 2020;85:35–40.

40. Hillmer AT, Nadim H, Devine L, Jatlow P, O’Malley SS. Acute alcohol consumption alters the peripheral cytokines IL-8 and TNF-α. Alcohol. 2020;85:95–99.

41. Brambilla F, Maggioni M. Blood levels of cytokines in elderly patients with major depressive disorder. Acta Psychiatrica Scandinavica. 1998;97(4):309–313.

42. Kagaya A, Kugaya A, Takebayashi M, et al. Plasma concentrations of interleukin-1β, interleukin-6, soluble interleukin-2 receptor and tumor necrosis factor α of depressed patients in Japan. Neuropsychobiology. 2001;43(2):59–62.

43. Zago A, Moreira P, Jansen K, et al. Alcohol use disorder and inflammatory cytokines in a population sample of young adults. Journal of Alcoholism & Drug Dependence. 2016:1–5.

44. Alfonso-Loeches S, Pascual-Lucas M, Blanco AM, Sanchez-Vera I, Guerri C. Pivotal role of TLR4 receptors in alcohol-induced neuroinflammation and brain damage. Journal of Neuroscience. 2010;30(24):8285–8295.

45. Alvarez Cooper I, Beecher K, Chehrehasa F, Belmer A, Bartlett SE. Tumour necrosis factor in neuroplasticity, neurogenesis and alcohol use disorder. Brain plasticity. 2020;6(1):47–66.

46. Gonzalez-Quintela A, Dominguez-Santalla M, Perez L, Vidal C, Lojo S, Barrio E. Influence of acute alcohol intake and alcohol withdrawal on circulating levels of IL-6, IL-8, IL-10 and IL-12. Cytokine. 2000;12(9):1437–1440.

47. Banks WA, Kastin AJ, Gutierrez EG. Penetration of interleukin-6 across the murine blood-brain barrier. Neurosci Lett. 1994;179(1-2):53–56.

48. Sun Y, Li N, Zhang J, et al. Enolase of Streptococcus Suis Serotype 2 Enhances Blood-Brain Barrier Permeability by Inducing IL-8 Release. Inflammation. 2016;39(2):718–726.

49. Heberlein A, Käser M, Lichtinghagen R, et al. TNF-α and IL-6 serum levels: neurobiological markers of alcohol consumption in alcohol-dependent patients? Alcohol. 2014;48(7):671–676.

50. Roberts AJ, Khom S, Bajo M, et al. Increased IL-6 expression in astrocytes is associated with emotionality, alterations in central amygdala GABAergic transmission, and excitability during alcohol withdrawal. Brain, behavior, and immunity. 2019;82:188–202.

51. Luo Y, He H, Zhang M, Huang X, Fan N. Altered serum levels of TNF-α, IL-6 and IL-18 in manic, depressive, mixed state of bipolar disorder patients. Psychiatry Research. 2016;244:19–23.

52. O’Connor JC, Lawson MA, Andre C, et al. Lipopolysaccharide-induced depressive-like behavior is mediated by indoleamine 2, 3-dioxygenase activation in mice. Molecular psychiatry. 2009;14(5):511–522.

53. Petrof EO, Khoruts A. From stool transplants to next-generation microbiota therapeutics. Gastroenterology. 2014;146(6):1573–1582.

54. Bajaj JS. Alcohol, liver disease and the gut microbiota. Nature Reviews Gastroenterology & Hepatology. 2019;16(4):235–246.

55. Barki N, Bolognini D, Jenkins L, Hudson B, Tobin A, Milligan G. P273 Chemogenetic analysis of how receptors for short chain fatty acids regulate the gut-brain axis. In: BMJ Publishing Group; 2021.

56. Kiefer F, Jahn H, Jaschinski M, et al. Leptin: a modulator of alcohol craving? Biological Psychiatry. 2001;49(9):782–787.

57. Sobhani I, Bado A, Vissuzaine C, et al. Leptin secretion and leptin receptor in the human stomach. Gut. 2000;47(2):178–183.

58. Ur E, Wilkinson DA, Morash BA, Wilkinson M. Leptin immunoreactivity is localized to neurons in rat brain. Neuroendocrinology. 2002;75(4):264–272.

59. Banks WA, Kastin AJ, Huang W, Jaspan JB, Maness LM. Leptin enters the brain by a saturable system independent of insulin. Peptides. 1996;17(2):305–311.

60. Wayne S, Neuhouser ML, Ulrich CM, et al. Association between alcohol intake and serum sex hormones and peptides differs by tamoxifen use in breast cancer survivors. Cancer Epidemiology and Prevention Biomarkers. 2008;17(11):3224–3232.

61. Mikolajczak P, Okulicz-Kozaryn I, Kamiñska E, et al. Effect of subchronic ethanol treatment on plasma and cerebrospinal fluid leptin levels in rats selectively bred for high and low alcohol preference. Polish journal of pharmacology. 2002;54(2):127–132.

62. Tan X, Sun X, Li Q, et al. Leptin deficiency contributes to the pathogenesis of alcoholic fatty liver disease in mice. The American journal of pathology. 2012;181(4):1279–1286.

63. Santolaria F, Pérez-Cejas A, Alemán M-R, et al. Low serum leptin levels and malnutrition in chronic alcohol misusers hospitalized by somatic complications. Alcohol and Alcoholism. 2003;38(1):60–66.

64. Röjdmark S, Calissendorff J, Brismar K. Alcohol ingestion decreases both diurnal and nocturnal secretion of leptin in healthy individuals. Clinical endocrinology. 2001;55(5):639–647.

65. Bouna-Pyrrou P, Muehle C, Kornhuber J, Weinland C, Lenz B. Body mass index and serum levels of soluble leptin receptor are sex-specifically related to alcohol binge drinking behavior. Psychoneuroendocrinology. 2021;127:105179.

66. Montague CT, Prins JB, Sanders L, Digby JE, O’Rahilly S. Depot-and sex-specific differences in human leptin mRNA expression: implications for the control of regional fat distribution. Diabetes. 1997;46(3):342–347.

67. Stangl BL, Vatsalya V, Zametkin MR, et al. Exposure-response relationships during free-access intravenous alcohol self-administration in nondependent drinkers: influence of alcohol expectancies and impulsivity. International journal of neuropsychopharmacology. 2017;20(1):31–39.

68. Airapetov M, Eresko S, Lebedev A, Bychkov E, Shabanov P. The role of Toll-like receptors in neurobiology of alcoholism. BioScience Trends. 2021;15(2):74–82.

69. Wang S-C, Chen Y-C, Chen S-J, Lee C-H, Cheng C-M. Alcohol addiction, gut microbiota, and alcoholism treatment: A review. International journal of molecular sciences. 2020;21(17):6413.

70. Jerlhag E. Gut-brain axis and addictive disorders: A review with focus on alcohol and drugs of abuse. Pharmacol Ther. 2019;196:1–14.

71. Ding J-H, Jin Z, Yang X-X, et al. Role of gut microbiota via the gut-liver-brain axis in digestive diseases. World Journal of Gastroenterology. 2020;26(40):6141.

72. Galland L. The gut microbiome and the brain. Journal of medicinal food. 2014;17(12):1261–1272.

73. Vatsalya V, Kong M, Marsano LM, et al. Interaction of heavy drinking patterns and depression severity predicts efficacy of quetiapine fumarate XR in lowering alcohol intake in alcohol use disorder patients. Substance abuse: research and treatment. 2020;14:1178221820955185.

74. Kurlawala Z, Vatsalya V. Heavy alcohol drinking associated akathisia and management with quetiapine XR in alcohol dependent patients. Journal of addiction. 2016;2016.

75. Agabio R, Colombo G. GABAB receptor ligands for the treatment of alcohol use disorder: preclinical and clinical evidence. Frontiers in neuroscience. 2014;8:140.

76. Kashem M, Šerý O, Pow D, Rowlands B, Rae C, Balcar V. Actions of alcohol in brain: Genetics, Metabolomics, GABA receptors, Proteomics and Glutamate Transporter GLAST/EAAT1. Current Molecular Pharmacology. 2020.

77. Tobin AB, Barki N, Bolognini D, et al. Chemogenetic analysis of how receptors for short chain fatty acids regulate the gut-brain axis. bioRxiv. 2020.

78. Bjørkhaug ST, Aanes H, Neupane SP, et al. Characterization of gut microbiota composition and functions in patients with chronic alcohol overconsumption. Gut microbes. 2019;10(6):663–675.

79. Mostafa H, Amin AM, Teh C-H, Arif NH, Ibrahim B. Plasma metabolic biomarkers for discriminating individuals with alcohol use disorders from social drinkers and alcohol-naive subjects. Journal of substance abuse treatment. 2017;77:1–5.

80. Russo R, Cristiano C, Avagliano C, et al. Gut-brain axis: role of lipids in the regulation of inflammation, pain and CNS diseases. Current medicinal chemistry. 2018;25(32):3930–3952.

81. Asano Y, Hiramoto T, Nishino R, et al. Critical role of gut microbiota in the production of biologically active, free catecholamines in the gut lumen of mice. American Journal of Physiology-Gastrointestinal and Liver Physiology. 2012;303(11):G1288–G1295.

82. Bercik P, Denou E, Collins J, et al. The intestinal microbiota affect central levels of brain-derived neurotropic factor and behavior in mice. Gastroenterology. 2011;141(2):599–609, 609.e591-593.

