## Supplement to GBA for "Illustration of a Novel Gut-Brain Axis of Alcohol Withdrawal, Withdrawal-Associated Depression, Craving and Alcohol-Severity Index in Alcohol Use Disorder Patients"

**Supplementary Material**

**Methods**

*Clinical and Research Data*

Forty-eight enrolled study participants were grouped categorically using the Clinical Institute Withdrawal Assessment of alcohol scale (CIWA) as clinically significant CIWA group (CS-CIWA [score >10] Gr. 1 [n=22]), and clinically-not significant group (NCS-CIWA [score≤10] Gr. 2 [n=26]) ^1^. We used ≥10 as eligibility criteria for a diagnosis based on mild and above [severity of withdrawal for identifying withdrawal status](http://www.regionstrauma.org/blogs/ciwa.pdf). Clinical data and blood samples were collected upon enrollment. Demographics (Age, Sex, Body Mass Index [BMI]) and recent drinking history information were also recorded at the time of study visit. CIWA ^1^ scores, MADRS ^2^ assessment, craving assessment (measured by the Penn Alcohol Craving Scale [PACS]) ^3^, 90-day Timeline followback ^4^ (TLFB90: TD90, NDD90, AvgD90, HDD90) for recent drinking profile, lifetime drinking history (LTDH) ^5^, and alcohol composite score (ACS, from the Addiction Severity Index assessment [ASI] ^6^) were also collected for analyses. Montgomery-Asberg Depression Rating Scale criteria for reference ranges were used ([Montgomery-Asberg Depression Rating Scale (MADRS) - MDCalc](https://www.mdcalc.com/montgomery-asberg-depression-rating-scale-madrs#evidence)) from previously reported publication ^2,7^. PACS was performed on a sub-set of 16 AUD study patients; and its association with the markers of gut-alterations, heavy drinking, and inflammation was assessed. PACS ≥6 was used for identifying drinking levels and the cytokine/ gut-permeability measures which might predict moderate to high craving ^8^.

Information on recent drinking and the patterns of drinking was collected from the Timeline Followback questionnaire ^4^. Measures were assessed for the past 90 days and included Total Drinks (TD90), Number of Drinking Days (NDD90), Number of Non-Drinking Days (NNDD90), Average Drinking per Drinking Days (AvgDPD90), and Heavy Drinking Days (HDD90). Chronic drinking (as the number of years) was assessed using the LTDH (in yrs.) questionnaire ^5^. We also used the “Controlling Nutritional Status Test” (CONUT) to establish the nutritional status ^9^ of our patients.

*Bodily Samples and Laboratory Testing*

Blood samples were collected at the time of the study visit for each enrolled patient. Blood samples were processed for plasma and frozen at -80° C until assayed. Plasma samples were analyzed for the pro-inflammatory cytokines (interleukin-6 [IL-6], interleukin-8 [IL-8], tumor necrosis factor- α [TNF-α], interleukin-1β [IL-1β] and monocyte chemoattractant protein-1 [MCP-1]) and adiponectin, and leptin by multianalyte chemiluminescent detection using Multiplex kits (Millipore, Billerica, MA, USA) on the Luminex (Luminex, Austin, TX, USA) platform based on the manufacturers’ instructions. Plasma LPS and LPS binding-protein(LBP), and +sCD14 were assessed using the Kinetic Chromogenic Limulus Amoebocyte Lysate Assay (Lonza, Walkersville, MD, USA) following the manufacturer’s instructions.

*Statistical Analyses*

Univariate factorial ANOVA was used to evaluate the differences in the demographic and drinking history markers, scores of CIWA and other alcohol-associated domains such as depression; gut-permeability measures, and levels of various cytokines. Univariate ANOVA was used to evaluate these differences by an additional discrete secondary factor of sex as between-group response and statistical interaction for sub-set assessments. Sex-based association effects within each sex were also evaluated as per the scope. Drinking history and other demographic factors were tested as confounders of inflammation in AUD. Linear regression analysis was used to characterize the association of drinking domains and drinking measures or along with the laboratory biomarkers. The laboratory markers of AUD domains, such as hormones and proinflammatory cytokines (adiponectin, IL-1β, IL-6, IL-8, TNF-α, MCP-1, leptin, and other pro-inflammatory cytokines and hormones) were tested for the within-group associations with the clinical and drinking markers; and with gut-permeability measures (LPS, LBP, sCD14) using the linear regression analysis (as univariate or multivariate independent variable models). Gut-brain axis model specific for candidate AUD domains of this study were constructed for withdrawal, depression, and craving incorporating the role of the immunological status. A predictive model on a sub-set of the study patients was developed for characterizing the gut-brain axis of craving based on the PACS score using a multivariate regression model with stepwise addition of variable/s contributing at various steps of gut-brain pathway. PACS score was also used as a factor to assess the prediction model of heavy drinking patterns and candidate gut-immune measures. SPSS 27.0 (IBM, Chicago, IL, USA) and Microsoft 365 Excel application (MS Corp, Redmond, WA, USA) were used for statistical analysis and data computation. Statistical significance was established at *p*≤0.05. Data are expressed as M ± SD (Mean ± standard deviation), unless otherwise noted specifically.

**Results**

**Alcohol Addiction Severity Index**

Alcohol Composite Score (ACS, another measure of alcohol drinking that describes the Addiction Severity Index) and HDD90 measures were significantly and positively associated (R²=0.318 at p≤0.001) as measure of heavy drinking consequential in severity due to alcohol drinking across all the patients (S. Fig 1a). ACS was also found to be significantly associated with the gut permeability markers, Lipopolysaccharide (LPS), and Lipid binding protein (LBP) respectively across all the patients (S. Fig 1b-1c).

**Discussion**

AUD patients show a corresponding positively related response by the degree of heavy drinking in recent active past and the severity index of alcohol drinking as reported by ACS ^6^. Our evaluation supports a pathological connection of the correspondingly increasing order of the severity index of alcohol drinking and LPS (an altered gut dysfunction response characterized by the alcohol associated endotoxemia ^10^). Elevated LBP with low ACS could indicate its enhancing role, albeit lower LBP at higher ACS could well be directed towards decreasing the biological activity of LPS, when LPS is in excess ^11^, as observed with a negative relationship in our findings. A valid questionnaire such as ACS based assessment could present a valuable link between the recent heavy drinking pattern and the gut-dysregulation that is encountered in the AUD patients.

**Supplementary Figure Legend**

Supplementary Figure Legend 1. Severity of alcohol drinking, and its associaiton with alcohol drinking pattern and gut dysregulation in all AUD patients. Fig. S1a. Heavy drinking days in the past 90-day (HDD90) showed high positive relationshsip with the alcohol composite acore (ACS). Fig. S1b. ACS showed significant positive associaiton with the lipopolysaccaride. Fig. S1c. ACS showed significant negative associaiton with the lipopolysaccaride-binding protein (LBP). Statistical significance was set at p<0.05.

**References**

1.	Sullivan JT, Sykora K, Schneiderman J, Naranjo CA, Sellers EM. Assessment of alcohol withdrawal: the revised clinical institute withdrawal assessment for alcohol scale (CIWA‐Ar). *British journal of addiction.* 1989;84(11):1353-1357.

2.	Müller MJ, Szegedi A, Wetzel H, Benkert O. Moderate and severe depression. Gradations for the Montgomery-Asberg Depression Rating Scale. *J Affect Disord.* 2000;60(2):137-140.

3.	Flannery B, Volpicelli J, Pettinati H. Psychometric properties of the Penn alcohol craving scale. *Alcoholism: Clinical and Experimental Research.* 1999;23(8):1289-1295.

4.	Sobell LC, Sobell MB. Timeline follow-back. In: *Measuring alcohol consumption.* Springer; 1992:41-72.

5.	Skinner HA, Sheu W-J. Reliability of alcohol use indices. The Lifetime Drinking History and the MAST. *Journal of studies on alcohol.* 1982;43(11):1157-1170.

6.	Rosen CS, Henson BR, Finney JW, Moos RH. Consistency of self‐administered and interview‐based Addiction Severity Index composite scores. *Addiction.* 2000;95(3):419-425.

7.	Montgomery SA, Åsberg M. A new depression scale designed to be sensitive to change. *The British journal of psychiatry.* 1979;134(4):382-389.

8.	Yoon G, Kim SW, Thuras P, Grant JE, Westermeyer J. Alcohol Craving in Outpatients with Alcohol Dependence: Rate and Clinical Correlates. *Journal of Studies on Alcohol.* 2006;67(5):770-777.

9.	Fukushima K, Ueno Y, Kawagishi N, et al. The nutritional index ‘CONUT’is useful for predicting long-term prognosis of patients with end-stage liver diseases. *The Tohoku journal of experimental medicine.* 2011;224(3):215-219.

10.	Kirpich IA, McClain CJ, Vatsalya V, et al. Liver injury and endotoxemia in male and female alcohol‐dependent individuals admitted to an alcohol treatment program. *Alcoholism: Clinical and Experimental Research.* 2017;41(4):747-757.

11.	Lamping N, Dettmer R, Schröder N, et al. LPS-binding protein protects mice from septic shock caused by LPS or gram-negative bacteria. *The Journal of clinical investigation.* 1998;101(10):2065-2071.
