## Supplementary figures and images for "Illustration of a Novel Gut-Brain Axis of Alcohol Withdrawal, Withdrawal-Associated Depression, Craving and Alcohol-Severity Index in Alcohol Use Disorder Patients"

### Supplement Fig. 1

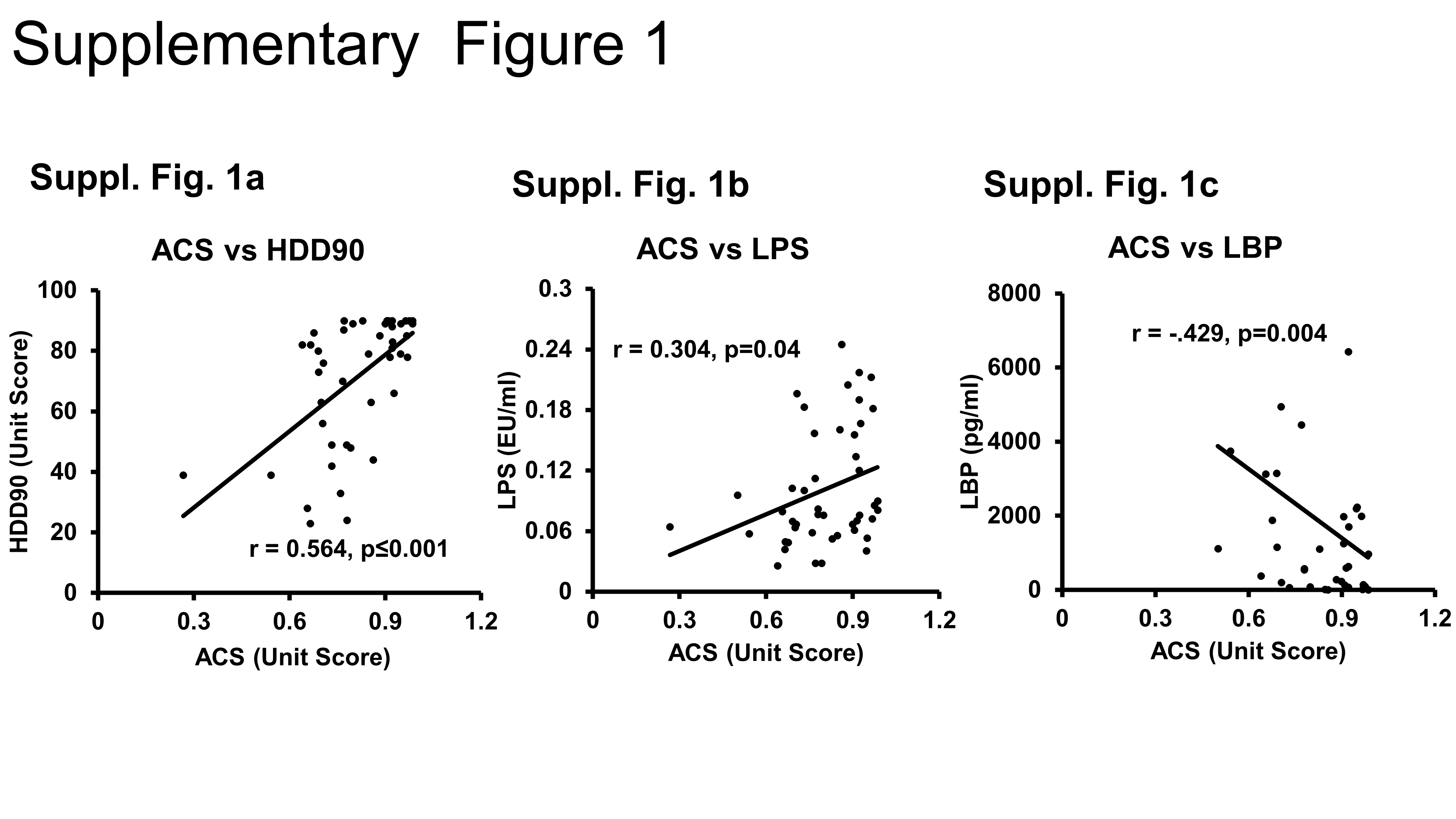
